# Estimating LFT and qPCR test-sensitivity over time since infection from a human challenge study

**DOI:** 10.1101/2022.10.18.22280274

**Authors:** Emma L Davis, T Deirdre Hollingsworth

## Abstract

Testing has been central to global policy throughout the SARS-CoV-2 pandemic. Understanding how test sensitivity changes after exposure is crucial for the interpretation of test outcomes and the design of testing-based interventions. Using data from a human challenge study, we derive temporal test sensitivity profiles for lateral flow tests (LFT), quantitative polymerase chain reaction (qPCR) tests and viable virus, measured by focus-forming assay (FFA). The median time to detectability was 2 days (throat swab) and 3 days (nasal) for qPCR and 4 days for LFT (both swabs), and there was strong positive correlation between first LFT positive and first FFA positive for both throat (p=0.00019) and nasal (p=0.00032) swabs, supporting the use of LFTs as a method of detecting infectiousness. Peak LFT sensitivity was 82.4% (67.0%-91.8%) for throat samples, occurring 6 days post-exposure and 93.3% (85.1%-98.0%) for nasal, 7 days post-exposure. These temporal profiles provide quantification of the mean behaviour of these tests and individual-based variability and could inform a framework for investigating future testing-based interventions.

## Main text

The control of COVID-19 through both home-based and lab-based testing has been a step-change in infection management globally, but ongoing uncertainties around the sensitivity of commonly used tests – including the temporal sensitivity profile – have sparked discussion around whether these tests are being used appropriately^1–3^. Using data from a human challenge study^4^ we derive a model for test sensitivity over time since exposure for three test methods: rapid lateral flow tests (LFTs); laboratory-based quantitative polymerase chain reaction (qPCR) tests; and viable virus, measured by focus-forming assay (FFA).

Testing forms a crucial part of the public health response to any infectious disease, enabling not only accurate diagnosis, but informing surveillance and the targeting of interventions. During the SARS-CoV-2 pandemic, testing has been used for a range of purposes, including the quarantine of infectious individuals, assessing trends in prevalence to support policy interventions such as movement restrictions and face-mask requirements, and to identify patients requiring pharmaceutical treatments.

As well as supporting public-health policy, the increasing availability of LFTs has paved the way for individual-level risk management around decisions such as participation in mass events or visiting at-risk relatives. Countries such as the UK, Germany, Spain, Canada and France have provided free access to self-test kits at various points across the pandemic, allowing unprecedented public access to rapid at-home testing. Although these policies have mostly been withdrawn, LFTs remain easily accessible at relatively low cost and are still an important tool for the individual. They also continue to be used by businesses and venues and may still have policy-relevant applications in the future.

Early estimates of low LFT sensitivity and specificity sparked concerns around the appropriateness of LFTs as a public health tool^1^, with sensitivity ranging from 34.1% to 88.1%^5^ and specificity from 92.4% to 100.0%^6^, although these estimates were demonstrated to vary with factors such as symptoms of infection^5^, the administration route^5^, and viral load – which changes over the course of infection^7^. An additional consideration has been the comparability of LFTs with other test types, such as qPCR, which has a longer lead time (often 24-48 hours) and is more expensive than LFTs but is widely considered the gold-standard for SARS-CoV-2 detection, with higher reported sensitivity and specificity^5^.

Retrospective analyses of household studies^8^, and hospital studies comparing LFT positivity and detection of infectious virus^9^, have highlighted that LFTs are potentially a good test for infectivity, but it is unclear how this changes over the course of an infection. Each of these studies highlight the importance of time since infection as an unobserved variable. Understanding the temporal dynamics of positivity is critical for designing nuanced interventions, such as twice-weekly testing for schools in the UK or test-to-release quarantine schemes. It is also a crucial parameter to include in transmission models which are used to inform policy design. Using data from a human challenge study^4^, we re-evaluate the temporal characteristic of LFTs, qPCR and FFA using survival analysis.

During the human challenge study 34 participants aged 18-29 years without evidence of previous infection or vaccination were inoculated with 10 TCID50 of a wild-type virus and then monitored, with 18 participants becoming infected. Individual-based daily LFT, qPCR and FFA test results for 14 days post-exposure were published in Supplementary Figure 1 alongside the study.

We analysed this data to estimate temporal sensitivity, by fitting time-to-event models (survival analysis) for the time until first positive test for LFT, FFA and qPCR (see Figure 1) and time to last positive test for LFT and FFA tests. These survival functions are combined to derive a temporal expression for virus detectability over time since exposure, scaled by the maximum sensitivity, which is calculated as the proportion of tests returning positive in the period between a participant’s first and last positive test (see Figure 2). In the supplementary information we provide detailed instructions for applying these models within epidemic modelling frameworks for future more detailed policy development.

**Figure 1:**
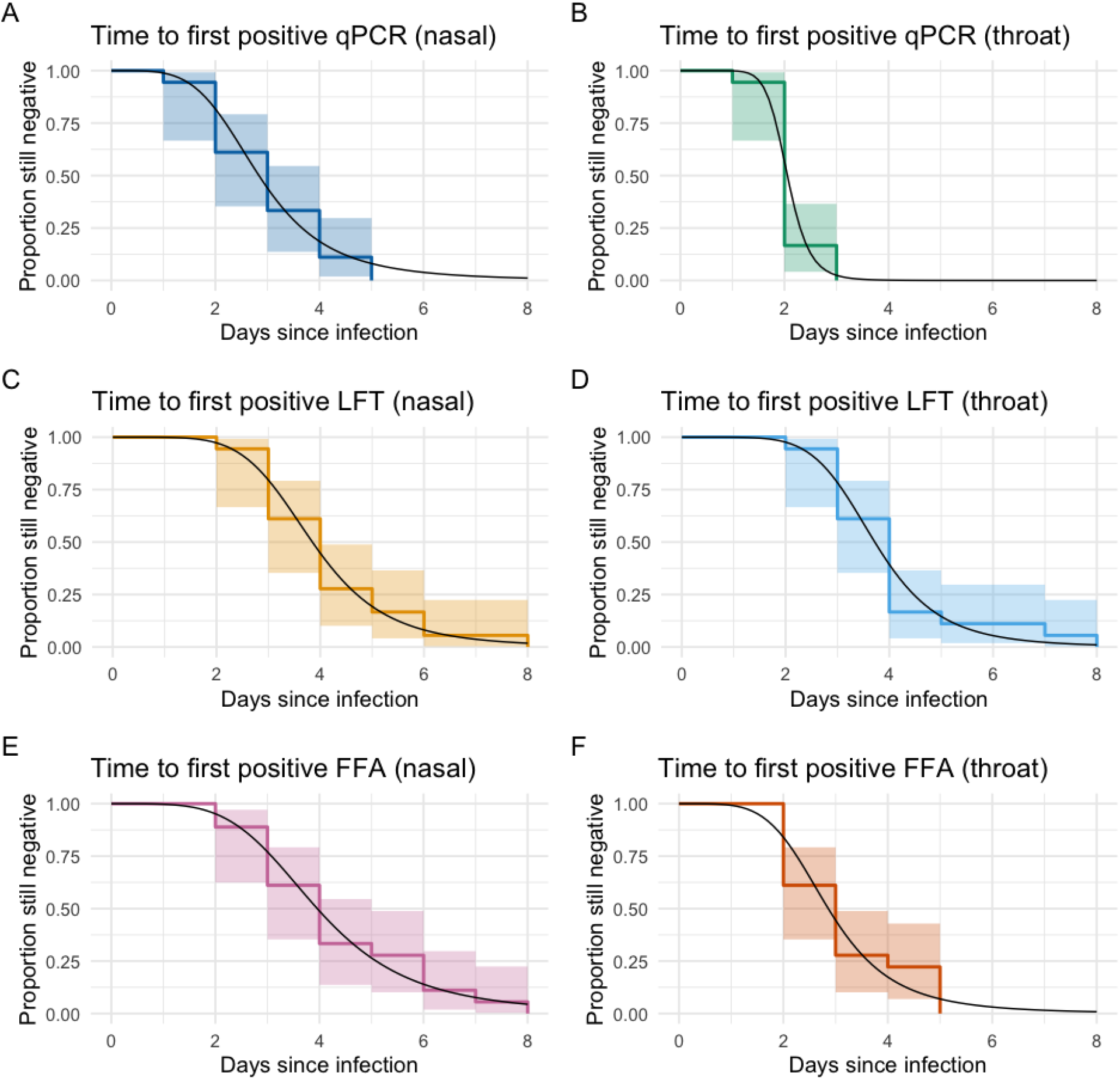
Survival curves for time to first positive test since infection across (from top to bottom) qPCR, LFT and FFA test types for nasal (left) and throat (right) swabs, fitted using the survival package in R version 4.0.3 assuming a log-logistic distribution. Shaded regions represent 95% log-log pointwise confidence intervals.

**Figure 2:**
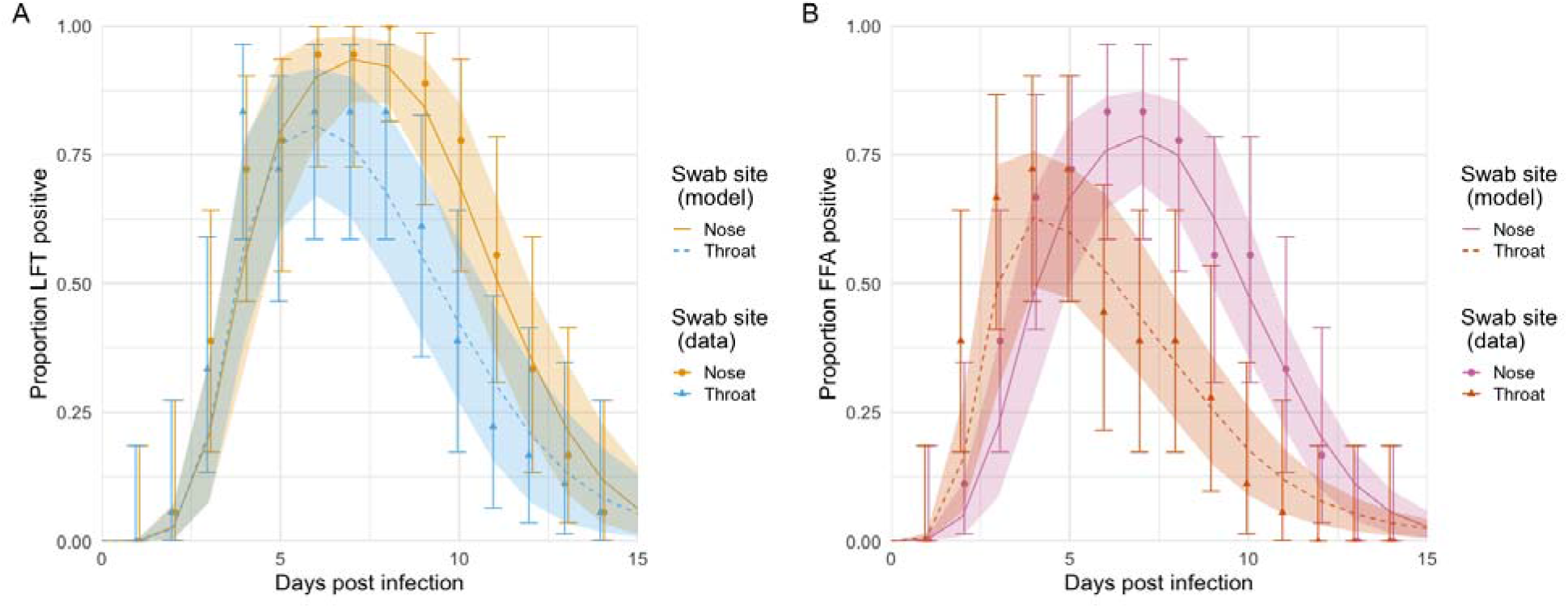
Probability of testing positive over time since infection for LFT (left) and FFA (right). Models are displayed as a solid line (nose swabs) or a dashed line (throat swabs), with shaded regions representing 95% confidence intervals, obtained via percentile bootstrap (10,000 resamples). Data points are displayed as circles (nose swabs) or triangles (throat swabs), with error bars representing 95% binomial confidence intervals.

The first parameter of interest is the time from infection to first testing positive. As expected, detection is earliest by qPCR, with throat tests providing positive results for all individuals by day 3 (Figure 1B, median 2 days, range 1-3 days). For nasal samples there is greater variability between individuals (Figure 1A, median 3 days, range 1-5 days). A student’s t-test demonstrates a significant difference (p=0.0056) between mean time to first qPCR positive by nasal and throat swabs.

The results for LFT tests are more similar across nasal and throat samples: the first individual testing positive on day 2 and all individuals positive by day 8 (median 4 days, Figures 1C and D) for both swab types, showing no significant difference between nasal and throat (p=0.82).

The first day on which viable virus was detected by FFA was on day 2, with all individuals exhibiting viable virus by at least one swab type by day 5 (Figure 1E and F). Although there was no significant individual-level correlation between timing of first positive across nasal and throat swab types (p=0.11), viable virus was detectable on average 1.17 days earlier (IQR 0.25-2.00 days) by throat swab than nasal swab and there was a significant difference in expected time to first positive (p=0.024).

There was strong positive correlation between first LFT positive and first FFA positive for both throat (p=0.00019) and nasal (p=0.00032) swabs, supporting the use of LFTs as a method of detecting infectiousness early in the infectious period. On average LFTs remain positive later than FFAs: 1.00 days via nasal swab (IQR 0.00-1.75 days) and 2.22 days via throat swab (IQR 0.25-4.00 days).

Due to the variability in individual measurements, the summary sensitivity of each test rises over the first few days of infection and then gradually declines over the following days (Figure 2). Since all individuals were still positive by qPCR on day 14, we present models for LFT and FFA only. The sensitivity of the LFT test peaked earlier and at a lower value (day 6, 82.4% (67.0%- 91.8%)) for throat samples than for nasal samples (day 7, 93.3% (85.1%-98.0%)). This temporal variation provides context for variable test sensitivity for studies which cannot control for time since infection.

Estimates of the sensitivity of FFA as a test of infection shows a similar pattern - sensitivity on nasal samples peaked later and higher (day 7, 78.7% (69.2%-87.4%)) than throat samples (day 4, 62.8% (49.3%-75.7%)). Since FFA is a potential surrogate for infectivity, this is an additional illustration that there is variability in infectivity over time and across individuals. The difference in sensitivity by swab location may also highlight that transmission by different routes may occur at different points in the days since infection, but the relationship between infectivity in the nose or throat and infectivity is still to be determined. However, these modelled estimates could be used to inform models of transmission over time since infection, which are currently informed by a limited number of serial interval datasets.

Our peak LFT sensitivity estimates (93.3% for nasal swabs and 82.4% for throat) are substantially higher than most non-temporal estimates in the literature^5,6^, due to most studies reporting mean estimates not including consideration of time since infection. Mean estimates (across the first 10 days since infection) given by the model are similarly low: 58.7% (nasal) and 48.0% (throat). This shows the importance of considering temporal variation in test sensitivity when discussing the appropriateness of testing-based interventions or analysing test results and data.

This is particularly important when considering the frequency of testing, for example to inform policy on repeat- or mass-testing. Additionally, interventions such as contact tracing are targeted to identify individuals earlier in their infection and have the benefit of a defined suspected exposure time, so an understanding of temporal sensitivity could allow for more targeted testing to maximise sensitivity^10^.

An understanding of the likelihood of false-negative results over time is also beneficial to the self-testing individual assessing their personal risk-level, for example making behavioural decisions after a potential period of increased exposure, and for the health practitioner when interpreting individual testing outcomes and advising patients.

Our modelled estimates of sensitivity over time since infection, based on the human challenge studies, provide important quantification of both mean behaviour of these tests and individual-based variability in detectability. These estimates are complementary to indirect estimates from larger scale studies, such as estimates of qPCR false-negative rate over time based on time relative to symptom onset^11,12^, and estimates of LFT detection of infectivity^9^.

Our estimates are based on a rare human challenge study, with a single variant from early in the pandemic and completely naive participants in a narrow age bracket (18-29 years). It is likely that these values have changed over time and will depend on individual infection history, demographic characteristics and vaccination status. Nevertheless, the functional forms of these relationships provide a framework for investigating sensitivity of future policies to these changes.

## Supporting information

Supplementary methods and results

## Data Availability

All data produced are available online at https://github.com/emmalouisedavis/temporaltestsensitivity

## Data Availability

The data used in this study is publicly available (https://doi.org/10.1038/s41591-022-01780-9) and is also provided at https://github.com/emmalouisedavis/temporaltestsensitivity

## Code Availability

All custom scripts to reproduce the analyses and figures presented in this article are available at https://github.com/emmalouisedavis/temporaltestsensitivity (release v1.0.0, DOI: 10.5281/zenodo.6977350).

## Acknowledgments

The authors would like to thank Daniel Pan, Thomas Crellen, Li Pi, Anna Borlase and Carl Whitfield for interesting discussions around the topics covered by this study.

