## Supplementary methods and results for "Estimating LFT and qPCR test-sensitivity over time since infection from a human challenge study"

**Section 1: Extended Methods**

The R code used in this study is available on GitHub and has been released on Zenodo, DOI: 10.5281/zenodo.6977350, URL: <https://github.com/emmalouisedavis/temporaltestsensitivity>

1.1 Human Challenge Data

The main dataset used in this study was taken from Supplementary Figure 1 of Killingley et al.^1^ The data represents a human challenge study where 34 participants aged 18-29 years without evidence of previous infection or vaccination were inoculated with 10 TCID50 of a wild-type virus and then monitored, with 18 participants becoming infected. Participants were tested by LFT, qPCR and FFA twice daily (nose and throat swabs), with daily test results reported in Supplementary Figure 1. Of the 18 infected participants, 6 received pre-emptive remdesivir (100mg intravenously for 5 days) once two consecutive 12-hourly nose or throat swabs tested positive by qPCR and 16 reported mild-to-moderate symptoms.

1.2 Survival Analysis

All survival analyses were done using the survival package and plots were created using the survminer package, both in Rstudio (R version 4.0.3). In these analyses “survival” was defined as “still testing negative” for time to first positive and “still testing positive, given tested positive at least once” for time to last positive.

Time to first positive analysis was done for all three test types, LFT, FFA and qPCR, and for nasal and throat swabs. Time to last positive analysis was done for LFT and FFA, nasal and throat swabs, but not for qPCR due to high levels of right censoring: 100% (18/18) for any nasal qPCR positive; 83.3% (15/18) for any throat qPCR positive; 72.2% (13/18) for strong nasal qPCR positive; and 77.8% (14/18) for strong throat qPCR positive.

First the survival::survdiff function was used to determine that neither treatment nor symptoms were significant factors for any of the tests across both swab types, so the data was pooled into one dataset for all further analyses.

Kaplan-Meier survival curves, $S(t)$, were generated using function survival::survfit and a log-logistic survival time distribution assumption was tested by plotting the relationship between log(time since exposure) and the log-failure odds.

Log-failure odds $=log(\frac{1-S(t)}{S(t)})$.

These plots are displayed in Figures S1 (time to first positive) and S2 (time to last positive), where a straight line indicates the log-logistic assumption is appropriate.

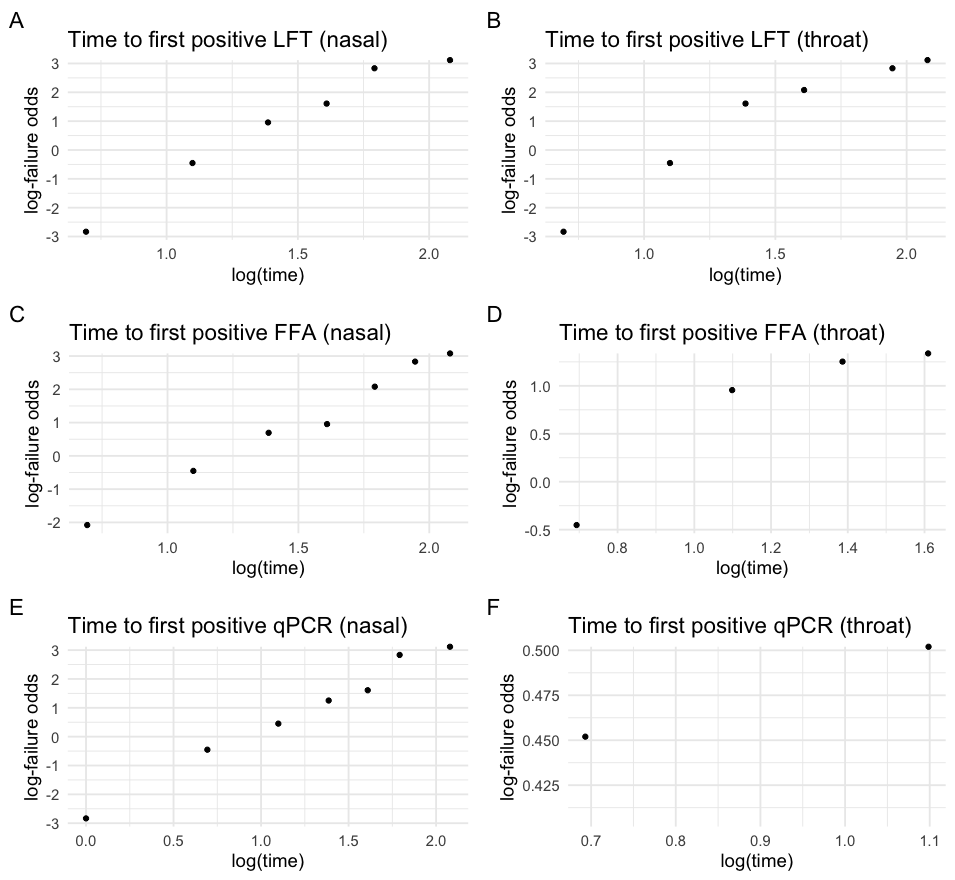

Figure S1: Log-failure odds plots testing a log-logistic survival time assumption for time to first positive test. From top: LFT, FFA, qPCR. Left: Nasal swabs. Right: Throat swabs.

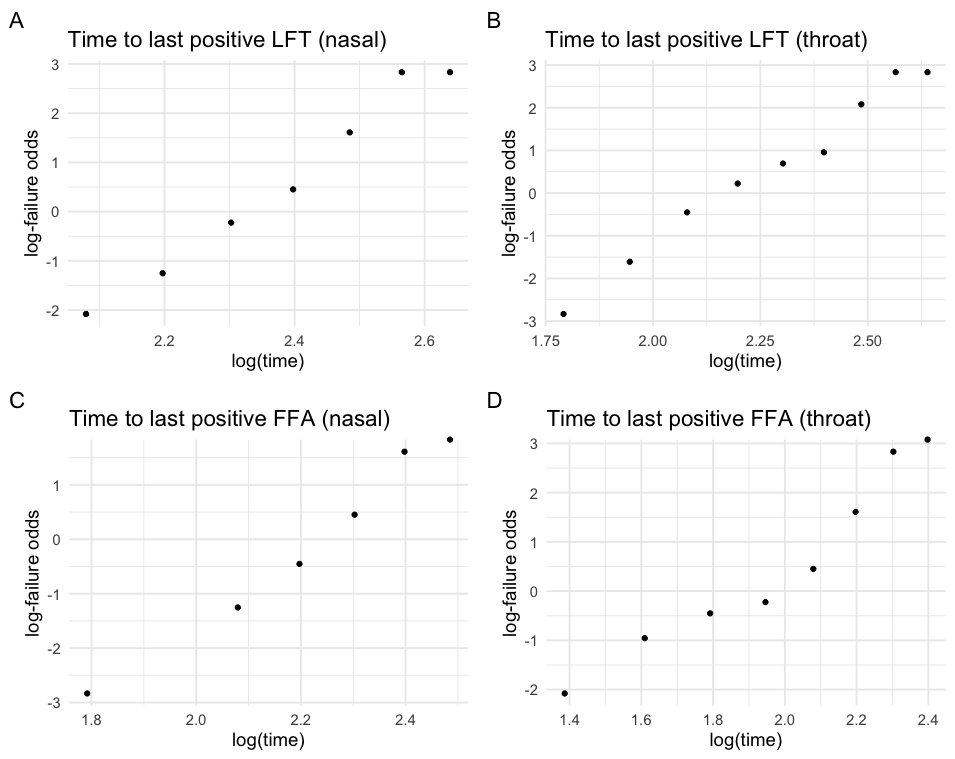

Figure S2: Log-failure odds plots testing a log-logistic survival time assumption for time to last positive test. From top: LFT, FFA, qPCR. Left: Nasal swabs. Right: Throat swabs.

Accelerated failure time (AFT) curves were then fitted using the survival::survreg function, assuming a log-logistic distribution. Fitted survival parameters are provided in Tables S1 (time to first positive) and S2 (time to last positive) for the following functional form of survival over time, $S(t)$:

$S(t)=\frac{1}{1+{\alpha t}^{\gamma}}$.

| Test | Swab | $\alpha$ (mean) | $\gamma$ (mean) |
| --- | --- | --- | --- |
| Lateral flow (LFT) | Nasal | $6.377\times10^{-4}$ | $5.459$ |
| Lateral flow (LFT) | Throat | $3.879\times10^{-4}$ | $5.979$ |
| Focus-forming assay (FFA) | Nasal | $2.450\times10^{-3}$ | $4.371$ |
| Focus-forming assay (FFA) | Throat | $7.621\times10^{-3}$ | $4.642$ |
| qPCR | Nasal | $2.362\times10^{-2}$ | $3.393$ |
| qPCR | Throat | $1.272\times10^{-3}$ | $8.022$ |

Table S1: Survival function parameters for time to first positive test, by test and swab type.

| Test | Swab | $\alpha$ (mean) | $\gamma$ (mean) |
| --- | --- | --- | --- |
| Lateral flow (LFT) | Nasal | $2.648\times10^{-11}$ | $10.244$ |
| Lateral flow (LFT) | Throat | $1.879\times10^{-7}$ | $6.927$ |
| Focus-forming assay (FFA) | Nasal | $8.391\times10^{-11}$ | $10.143$ |
| Focus-forming assay (FFA) | Throat | $2.933\times10^{-5}$ | $5.278$ |

Table S2: Survival function parameters for time to last positive test, by test and swab type.

1.3 Temporal Model of Test Sensitivity

The temporal model of test sensitivity over time since exposure was constructed from detectability profiles (based on the survival functions for time to first and last positive test) and estimates of test sensitivity during the detectable period, to give separate models for LFT and FFA sensitivity, separated by swab-type (nasal or throat).

Test sensitivity during the detectable period was calculated as the proportion of positive test results between each participant’s first positive and last positive test, with values displayed in Table S3. This effectively works as an upper bound on sensitivity for each test type.

| **Test** | **Swab** | **Sensitivity: mean (95% CIs)** |
| --- | --- | --- |
| Lateral flow (LFT) | Nasal | 98.1% (93.2% - 99.8%) |
| Lateral flow (LFT) | Throat | 87.1% (78.0% - 93.3%) |
| Focus-forming assay (FFA) | Nasal | 86.6% (77.3% - 93.1%) |
| Focus-forming assay (FFA) | Throat | 66.7% (53.3% - 78.3%) |
| qPCR | Nasal | 98.8% (95.6% - 99.9%) |
| qPCR | Throat | 92.9% (88.2% - 96.2%) |

Table S3: Calculated mean estimates of test sensitivity during the detectable period by test and swab type with 95% binomial confidence intervals.

The probability an individual tests positive at time $t$, $P(t)$, is then the sum of the probability that $t$ is the time of the first positive test, the probability that $t$ is between the first and the last positive test, scaled by sensitivity during the detectable period, $\phi$, and the probability that $t$ is the time of the last positive test.

$P(t\mathbb{)=P[}X=t]+\phi\mathbb{P[}X<t<Z\mathbb{]+P[}Z=t]$, where $X<Z$.

$X>0$ is the time of the first positive test, $Z$ is the time of the last positive test, and

$\mathbb{P[}X=t]=S_{X}(t-1)-S_{X}(t)==\frac{1}{1+\alpha_{X}({t-1)}^{Y_{X}}}-\frac{1}{1+\alpha_{X}t^{Y_{X}}}$,

$\mathbb{P[}X<t<Z] = \sum_{n=1}^{t-1} \mathbb{P[}X=n]\frac{S_{Z}(t)}{S_{Z}(n)}$,

$\mathbb{P[}Z=t] = \sum_{n=1}^{t-1} \mathbb{P[}Z=t\mathbb{]P[}X=n]$.

1.4 Calculating Confidence Intervals

Confidence intervals for the temporal positivity curves were calculated using the percentile bootstrap case-resampling method, which involved resampling the participants with replacement to generate 10,000 resamples of the same sample size as the original data (18 participants). Survival curves and detectable sensitivities were then calculated independently, following the same method as listed above, for each of the 10,000 resamples to give temporal positivity estimates. 95% confidence intervals were obtained from the 2.5th and 97.5th percentiles of the outputs at each time point.

**Section 2: Extended Results**

2.1 Time to Last Positive

Figure S3 shows the survival curves for time from exposure to last positive test for LFT and FFA tests, nasal and throat swabs. For LFTs the median time from exposure to the last positive test was 11 days (95% CI: 10-12) for nasal swabs and 9 days (95% CI: 8-11) for throat swabs. In comparison, the last positive FFA test occurred one day earlier on average: 10 days (95% CI: 9-11, nasal) and 8 days (95% CI: 5-9, throat). All participants were FFA negative by day 13, indicating that viable virus was not detectable after day 12, but 3 of 18 (16.7%) tested positive by LFT on or after day 13 (2 by nasal and throat swab, 1 by nasal swab only), and one individual was still testing positive by both nasal and throat swab on day 14, the last day recorded in the dataset.

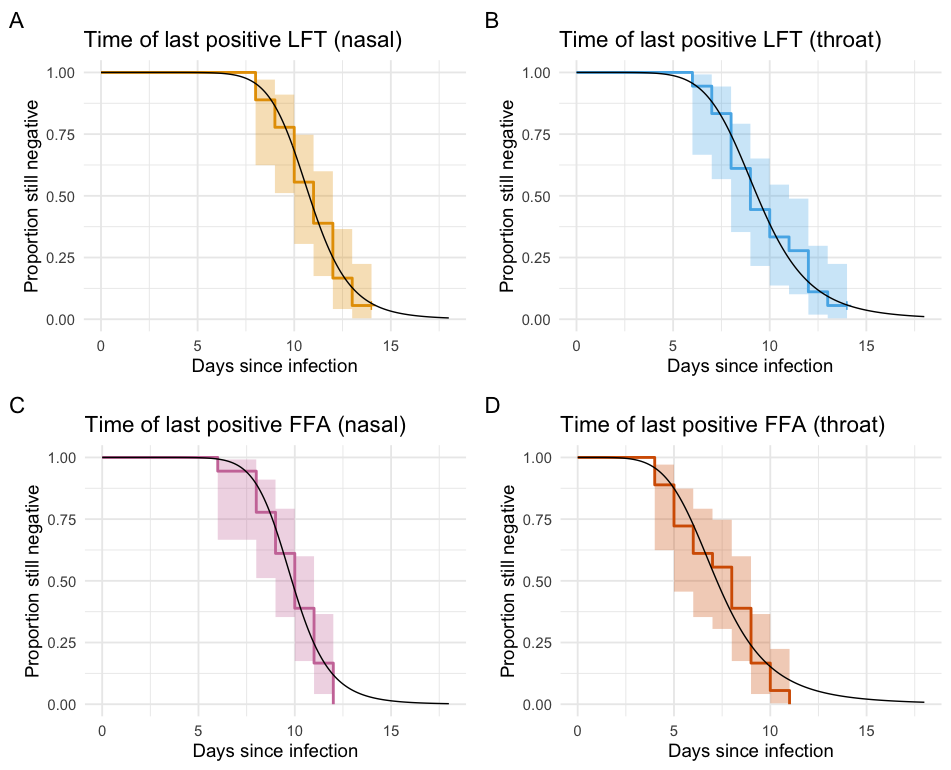

Figure S3: Survival curves for time to last positive test since infection across LFT (top) and FFA (bottom) test types for nasal (left) and throat (right) swabs, fitted using the survival package in R version 4.0.3 assuming a log-logistic distribution. Shaded regions represent 95% log-log pointwise confidence intervals.

2.2 Time Infectious

The period from first LFT positive to last FFA positive (Figure S4) could be considered as a proxy for the detectable period of infectivity, with first LFT positive representing the first time an individual might detect infection via self-test and the last FFA positive representing the last time at which viable virus is present, a sign of reduced infectiousness beyond this point.

The median time from first LFT positive to last FFA positive is 4.5 days (95% CI: 4-5) for nasal swabs and 4 days (95% CI: 3-4) for throat swabs. These estimates, and the narrow confidence intervals, indicate that may only be a short window of time in which individuals may test positive via LFT and still be shedding substantial quantities of infectious material.

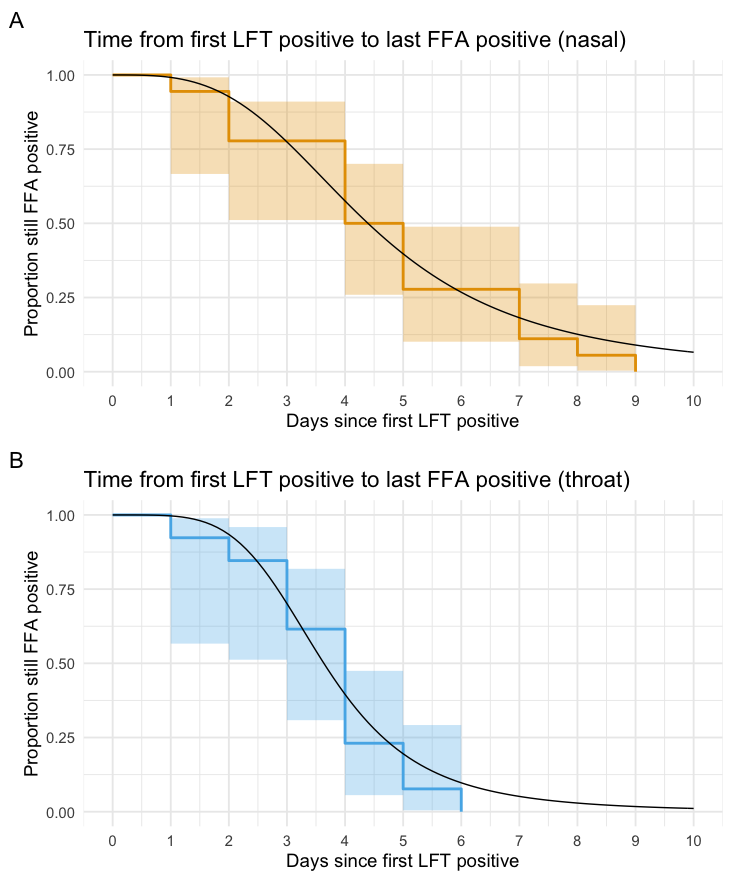

Figure S4: Survival curves for time from first LFT positive to last FFA positive test for nasal (top) and throat (bottom) swabs, fitted using the survival package in R version 4.0.3 assuming a log-logistic distribution. Shaded regions represent 95% log-log pointwise confidence intervals.

**Section 3: Additional qPCR Data**

3.1 Additional qPCR Data

The human challenge study from where the 14 day post-exposure day was sourced also reported additional qPCR data in Extended Data Figures 4 and 5. This data was presented in the form of a separate time series graph for each of the 18 participants and provides an additional 1-5 days of twice-daily testing per participant. We extracted this data using WebPlotDigitizer (<https://automeris.io/WebPlotDigitizer/>) and aligned this with the previously described daily testing data for days 1-14 to ensure consistent timing of tests across the combined dataset.

3.2 Additional qPCR Results

Following the steps outlined in Sections 1.2-1.4, we used this additional data to calculate time of last positive qPCR (split by strong/any positive and nasal/throat swab, see Figures S5 and S6) and temporal qPCR positivity over time (Figure S7).

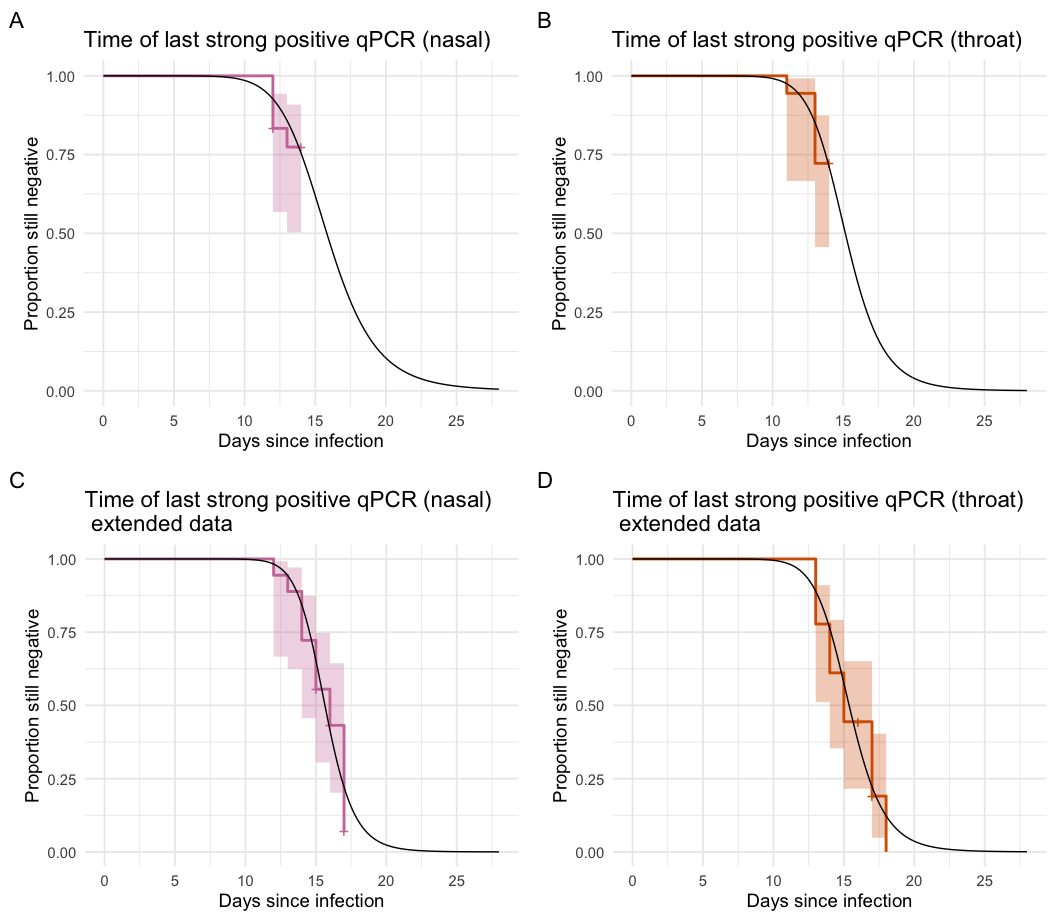

Figure S5: Survival curves for time to last strong positive qPCR test since infection for nasal (left) and throat (right) swabs, fitted using the survival package in R version 4.0.3 assuming a log-logistic distribution. Top row: Data used in main text (days 1-14). Bottom row: Extended data including days (15-19). Shaded regions represent 95% log-log pointwise confidence intervals.

Considering only strong qPCR positive test results, it was possible to get survival curves for time to last positive using just the data discussed in the main text (days 1-14, Figure S5: Top row), but there was still substantial censoring (14/18 participants by nasal swab and 13/18 participants by throat swab). However, including the additional data for days 15-19 (Figure S5: Bottom row) brought this to only 3/18 participants still testing strongly qPCR positive at the point of the last measurement for both swab types.

Median time from exposure to the last strong qPCR positive was 16 days (95% CI: 14-17) for nasal swabs, an average of 6 days later than the last nasal FFA positive, and 15 days (95% CI: 14-17) for throat swabs, an average of 7 days later than the last throat FFA positive. This reflects common knowledge that individuals are likely to be positive on qPCR for a substantial period after they are no longer infectious.

Figure S6 shows survival curves for the time to last positive qPCR, including strong and weak positive results and the additional data. Median time from exposure to the last qPCR positive was 18 days (95% lower confidence interval (LCI): 16) for nasal swabs, but the upper 95% confidence interval (UCI) was undefined due to censoring. For throat swabs, both the median and 95% UCI were undefined, due to censoring of more than 50% of the participants; the 95% LCI was 15 days.

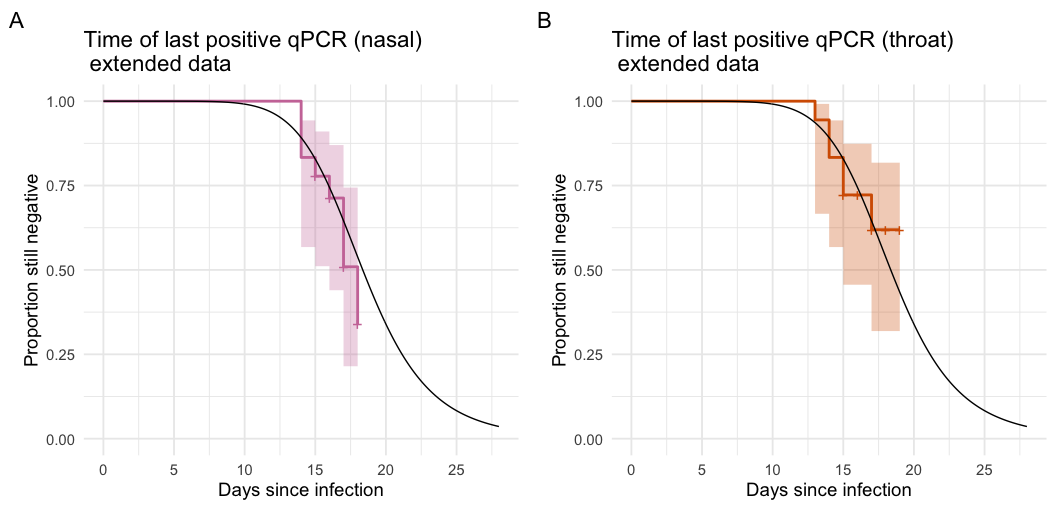

Figure S6: Survival curves for time to last positive (including weak positive) qPCR test since infection for nasal (left) and throat (right) swabs, fitted to the extended data including days (15-19) using the survival package in R version 4.0.3 assuming a log-logistic distribution. Shaded regions represent 95% log-log pointwise confidence intervals.

The utility of the additional data is demonstrated in the reduced width of the confidence intervals for temporal qPCR positivity over time since exposure (Figure S7) in panel B (strong positive only) compared to the model using the original data (panel A, strong positive only). It also allows us to consider the temporal probability of any qPCR positive, including weak positives (panel C).

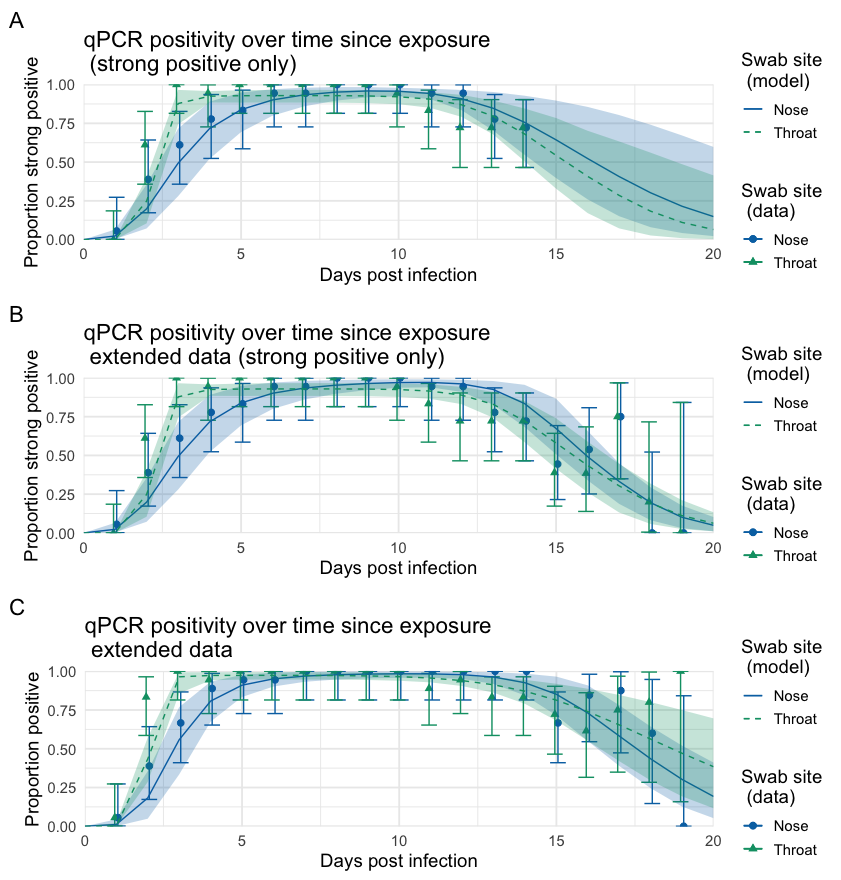

Figure S7: Probability of testing positive over time since infection for qPCR. Top: Original data (days 1-14), strong positive only. Middle: Extended data including days 15-19, strong positive only. Bottom: Extended data including days 15-19, any positive (including weak positive). Models are displayed as a solid line (nose swabs) or a dashed line (throat swabs), with shaded regions representing 95% confidence intervals, obtained via percentile bootstrap (10,000 resamples). Data points are displayed as circles (nose swabs) or triangles (throat swabs), with error bars representing 95% binomial confidence intervals.

For nasal swabs maximum sensitivity occurs on day 9 for the original model (95.9% (91.8%-98.4%), strong positive only) and on day 11 for strong qPCR positive (97.1% (94.0%-99.1%)) and day 10 for any qPCR positive (98.4% (95.8%-99.9%)) in the additional data model. For throat swabs maximum strong-positive sensitivity is 92.9% (88.4%-96.6%) on day 7 for both the original model and the additional data model and is 98.4% (93.9%-98.8%) on day 10 for all positives, including weak positives, in the additional data model.

**Section 4: Additional LFT Sensitivity Consideration**

4.1 Possible Factors

As mentioned in the main text, LFT sensitivity has been shown to vary by variables such as symptom presentation and swab method. In a review study Dinnes et al.^2^ reported the relative sensitivities associated with some of these variables, which can be summarised as a scaling factor relative to the ideal scenario (see Table S4). For the purpose of this analysis, it is assumed that participants in the human challenge study used in the model were swabbed by lab experts and, as they all reported at least one symptom across the study period, therefore represent measurements taken in the ideal scenario.

| Scenario | Scaling factor |
| --- | --- |
| Reference scenario: Symptomatic, swab by lab expert | 1 |
| Asymptomatic | 0.807 |
| Swab by NHS health worker | 0.888 |
| Self-swab | 0.730 |

Table S4: Scaling factors for asymptomatic cases and different swab administration routes.

4.2 Results

Figure S8 shows nasal LFT positivity since time of exposure for symptomatic versus asymptomatic infections (left, A) and difference swab administration routes (right, B).

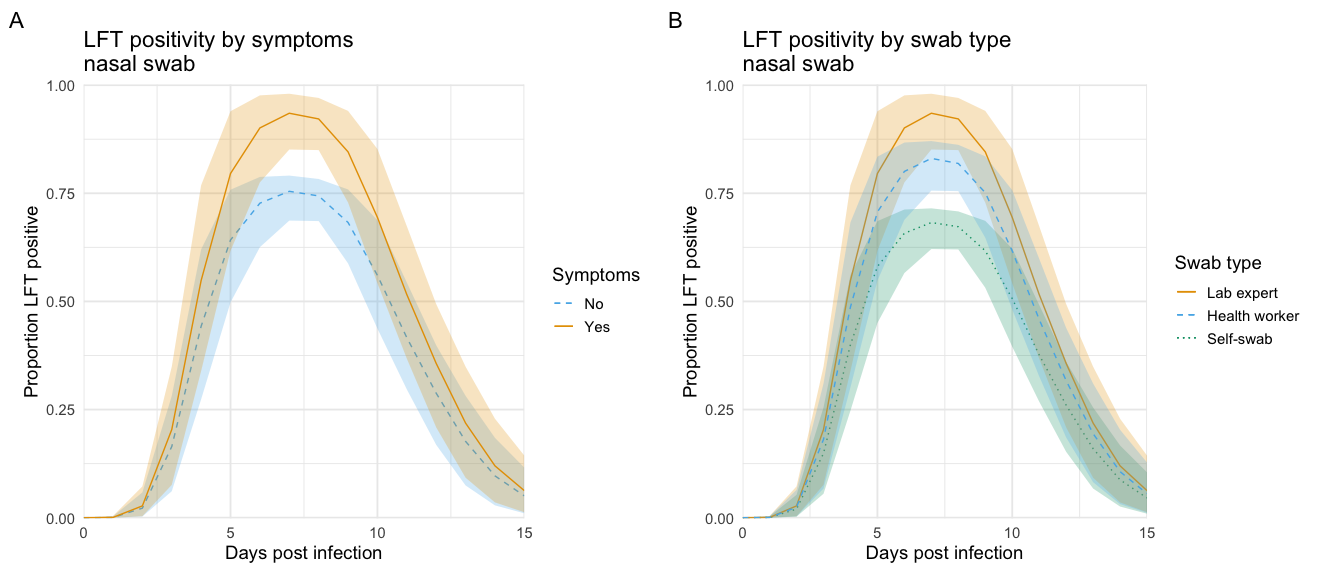

Figure S8: Probability of testing positive over time since infection for LFT using a nasal swab. Left: Scaled by symptoms yes (orange, solid) or no (blue, dashed). Right: Scaled by swab method: lab expert (orange, solid), health worker (blue, dashed), self-swab (green, dotted). Shaded regions representing 95% confidence intervals, obtained via percentile bootstrap (10,000 resamples).
